# How Images of Food Become Cravingly Salient in Obesity

**DOI:** 10.1101/2023.03.09.23287025

**Authors:** F. Devoto, A. Ferrulli, G. Banfi, L. Luzi, L. Zapparoli, E. Paulesu

## Abstract

**Background:** Obesity is characterized by neurofunctional alterations within the mesocorticolimbic reward system, a brain network that originates from the midbrain ventral tegmental area (VTA). Here we tested the hypothesis that this system might be misconfigured in obesity, reflecting a bias for food-related visual stimuli and a predisposition for craving.

**Methods:** A group of normal weight and a group of obese patients underwent a resting-state fMRI scan and the assessment of impulsivity, food craving, appetite, and implicit bias for food and non-food pictures. The VTA was used as a seed to map, for each participant, the strength of its functional connections with the rest of the brain. We then computed the difference of such maps between the groups and performed brain-behavior correlations.

**Results:** Obese patients showed hyper-connectivity of the VTA with part of ventral occipitotemporal cortices (vOTC) recently found to be specialized for food images; there was also a hypo-connectivity with the left inferior frontal gyrus (IFG), devoted to cognitive control. The VTA-left vOTC connectivity was positively associated with food craving and bias for high-calories food; a reversed such correlation was seen for the IFG.

**Interpretation:** These findings reveal that, in obesity, food-related visual stimuli become cravingly salient through an imbalanced connectivity of the reward system with sensory-specific regions and the frontal cortex involved in cognitive control.

## 1. Introduction

The worldwide prevalence of obesity is raising, casting serious concerns on its secondary consequences, such as musculoskeletal disorders, cardiovascular diseases, and diabetes, that increase the risk of mortality.^1^ Obesity is defined as excessive fat accumulation that may impair health, and the Body Mass Index (BMI) is commonly used to classify overweight (≥25 kg/m^2^) and obesity (≥30 kg/m^2^) in adults. Recently, cumulative evidence in neuroscience established the role of altered mesocorticolimbic activity and connectivity in the pathophysiology of obesity and substance use disorder, hinting to new potential clinical interventions.^2^

The mesocorticolimbic or reward circuitry originates from the ventral tegmental area (VTA) of the midbrain, a region that plays a pivotal role in the regulation of motivated behaviors.^3^ However, most human functional connectivity studies on obesity focused on major VTA efferents and afferents as regions-of-interest (ROIs) to map the functional connectome (e.g., nucleus accumbens, amygdala, anterior cingulate cortex, hypothalamus), leaving the possibility that the observed differences do not strictly pertain to neuroadaptations occurring at the roots of the reward system.^4^

Here, we characterized resting-state functional connectivity (rsFC) of the VTA in healthy weight and adults with obesity, to investigate whether obesity is associated with altered mesocorticolimbic connectivity. To validate the functional meaning of the observed rsFC patterns with respect to food cognition and behavior, we collected measures of impulsivity, food craving, appetite, and cognitive bias for food. In what follows we first describe the anatomy of VTA, its functional neuroanatomy, and its role in motivated behaviors. Then, we focus on previous evidence on patients with obesity suggesting the existence of an imbalance between brain networks involved in the processing of hedonic, homeostatic, and motivational aspects of food and those involved in higher-order cognitive processes. Finally, we spell out the specific aims of our study, the alternative hypotheses, and the expected results.

### 1.1. Functional neuroanatomy of the Ventral Tegmental Area

From the neurochemical point of view, 50-80% of VTA cells consists of dopamine neurons, while the remaining cells consist of subpopulations of GABAergic, glutamatergic, and combinatorial neurons^5,6^. Dopamine neurons exhibit reciprocal projections with subcortical and limbic structures like the hypothalamus and the ventral striatum (including the nucleus accumbens) - involved in appetite control, reward, and motivation - and the amygdala, hippocampus, and anterior insula - involved in incentive salience and memory.^7,8^ Fiber tracking studies also documented far reaching connections from the VTA to prefrontal regions, including its medial (anterior cingulate (ACC), ventromedial prefrontal cortex (vmPFC)) and lateral subdivisions (dorsolateral prefrontal cortex (dlPFC), lateral orbitofrontal cortex) - areas associated with reward evaluation, cognitive control, and voluntary goal-directed, as opposed to habitual, behavior^9,10^. rsFC studies in humans^11^ have also observed functional connections of the VTA with occipitotemporal and parietal cortices, suggesting different functional anatomical pathways through which the VTA can contribute to motivational processing.

### 1.2. Brain abnormalities in obese individuals: evidence from task-based and rsFC fMRI studies

Human neuroimaging studies highlighted two main neurocognitive hallmarks of obesity, both of which are thought to contribute to food craving: (i) increased reactivity of the reward system to food stimuli, linked to greater incentive salience attribution to food cues and to habitual responding; (ii) decreased involvement of prefrontal circuits, associated with lower cognitive control and to goal-directed behavior.

Compared with healthy weight controls, patients with obesity displayed greater activity of midbrain, subcortical (ventral striatum, amygdala) and medial prefrontal areas (ACC, vmPFC) in response to (high-calories) food cues, consistent with ongoing incentive motivational processes^12,13^. This reactivity of the reward system to food cues seems to be accompanied by a diminished recruitment of prefrontal areas when attempting to actively inhibit food cravings or behavioral responses to food cues, pointing to an imbalance between brain networks involved in motivation and those involved in higher-order cognitive processes.^14,15^ In a previous meta-analysis, we observed that obesity is associated with decreased activity of the ventral tegmental region and thalamus in response to food cues (consistent with a reward deficit theory of obesity), in spite of increased ventral striatal activity that persists in the satiety state, dovetailing with the incentive sensitization theory of addiction.^13,16-18^ We recently established that these functional alterations are complemented by overlapping structural abnormalities that extend to the vmPFC and ACC, reinforcing the notion that systems involved in motivation and those supporting cognitive control and decision-making are both involved in the pathophysiology of obesity.^19^

Results from rsFC studies suggest that the altered crosstalk between reward and cognitive control networks also holds without exogenous stimulation: patients with obesity exhibit increased rsFC between key nodes involved in homeostatic regulation (hypothalamus) and salience and reward processing (insula, ventral striatum), possibly reflecting incentive motivational processes that override homeostatic signals, leading to hedonic overeating.^20,21^ Decreased functional coupling between the amygdala and the vmPFC and between the hypothalamus and the dorsal ACC has also been reported, highlighting the contribution of frontostriatal circuits to the eating behavior.^22^

### 1.3. Aims of the study

In this seed-based rsFC study, we tested two key hypotheses: first, obesity is linked to altered rsFC of the VTA; second, the functional connectivity patterns of the VTA are associated with food craving and food-related cognitive processes.

To these aims, we mapped the whole-brain rsFC of the VTA in healthy weight and adults with obesity in a mild food-deprived state (4-5 hours) and complemented our neurofunctional measures with the assessment of impulsivity, food craving, and appetite. We also examined the specificity of the rsFC patterns with respect to food-related cognitive processes by assessing the cognitive bias for food and for non-food items.^23^

In keeping with the role of the mesocorticolimbic circuit in incentive salience and motivation, we expected patients with obesity to exhibit increased connectivity of the VTA with ventral striatal and limbic areas, and decreased connectivity with prefrontal regions involved in cognitive control. We hypothesized that these alterations may contribute to higher impulsivity and food craving in obesity, and that they would be specifically associated with the processing of food-related stimuli.

## 2. Material and Methods

### 2.1. Participants

Twenty-three healthy weight individuals, without any prior or current neurological or psychiatric disorder, and twenty-four otherwise healthy obese patients, matched for age, gender, and education were recruited for the study. Patients with obesity referred to the Endocrinology and Metabolic Diseases outpatient clinic at IRCCS Policlinico San Donato, for overweight/obesity treatment, and were part of a larger randomized, double-blinded, placebo-controlled clinical trial designed to study the effects of a deep repetitive Transcranial Magnetic Stimulation (deep rTMS) treatment aimed at reducing body weight and food craving^24^ (see^25^ for a description of the inclusion and exclusion criteria for participants with obesity). This study was performed prior to the beginning of the deep rTMS treatment, it was conducted in accordance with the 1964 Helsinki declaration, and it received approval from the local institutional review board (Ethics Committee of San Raffaele Hospital, Milan, Italy). All participants provided written informed consent before participating in any of the study procedures. One obese participant was excluded due to an incidentaloma (a meningioma located in the basal forebrain), resulting in twenty-three obese patients in the final sample. The implicit association test (IAT) data, measuring bias for high-calories food, from two patients were lost due to technical problems during response recording, resulting in 21 obese patients included in the analysis of IAT data. Demographic characteristics of the sample are reported in Table 1.

**Table 1.** Demographic characteristics of the sample.

|  | **Healthy weight**<br>**(n = 23)** | **Obese**<br>**(n = 23)** |  |
| --- | --- | --- | --- |
|  | **Mean (SD)** | **Mean (SD)** |  |
| Gender (M/F) | 7/16 | 6/17 | $\chi^2 = 0.11, p = 0.74$ |
| Age | 46 (11.5) | 51.4 (9.3) | $U = 186, p = 0.08$ |
| Education | 14.9 (3) | 14 (3.2) | $U = 225, p = 0.37$ |
| BMI (kg/m<sup>2</sup>) | 23.2 (1.5) | 36 (3.5) | $t(44) = -16.19, p < 0.001$ |
| Time from last meal (minutes) | 252 (40.5) | 300.6 (132.4) | $U = 187, p = 0.09$ |

### 2.2. Self-report data

#### Impulsivity

Upon arrival to the hospital, participants were asked to report the time of their last meal, and impulsivity was assessed with the Barratt Impulsiveness Scale-11 (BIS-11^26^), a 30-item self-reported questionnaire assessing different facets of impulsivity.

#### Food craving

Just prior to the functional Magnetic Resonance Imaging session, food craving was evaluated with the Food Craving Questionnaire (FCQ) Trait (FCQ-T) and State (FCQ-S) versions, a self-report inventory that investigates multiple dimensions of food craving.^27^

#### Hunger

Hunger was evaluated before fMRI with a 100 mm Visual Analogue Scales (VAS) ranging from 0 (not hungry at all) to 100 (extremely hungry).

#### Implicit bias for food

The implicit bias for food was assessed with the Implicit Association Test (IAT)^23^. The IAT is a categorization task during which participants must classify, as rapidly and accurately as possible, a set of pictures and words belonging to opposite (positive vs. negative) categories by means of button presses. It assumes that reaction times of sorting will be slower and less accurate during incongruent trials, when opposite categories must be classified together (e.g., positive pictures and negative words), compared to the congruent trials (e.g., positive pictures and positive words). This increased latency is thought to reflect the strength of association, and it is typically summarized by a score called D600^28^. Two versions of the IAT were administered: one featuring food stimuli (food: liked high-calories foods vs. disliked low-calories foods), and one featuring stimuli unrelated to food (non-food: flowers vs. weapons) (see the Supplementary Materials for further details about the tasks’ structure and stimuli). For each participant, we computed the D600 score for the food and non-food versions of the IAT to obtain a single measure of food-related and food-unrelated processing.

### 2.3. Neuroimaging data

#### fMRI data acquisition

MRI scans were performed using a 1.5 Siemens Avanto scanner, endowed with echo-planar hardware for imaging. Technical features of MRI and details on the preprocessing pipeline are reported extensively in the Supplementary Materials.

#### Seed-based functional connectivity

The left and right VTA were extracted as a single volume of interest from the Automatic Anatomical Labeling 3 (AAL3) template (https://www.gin.cnrs.fr/en/tools/aal/) by means of the MRIcron software, and used as a volume of interest (VOI) for a seed-based functional connectivity analysis.^29^ At first level, Conn computes bivariate correlations between the average BOLD signal within the VOI (from unsmoothed volumes) and every other voxel in the brain. The result is a rsFC map for each subject: these were entered into a second-level random-effect analysis conforming to an independent samples t-test.

### 2.4. Data analysis

#### Analytical strategy of self-report and neuroimaging data

The self-reported data were analyzed by means of the software Jamovi (Version 2.3.18). The Shapiro-Wilk test was used to assess the assumption of normality for the variables of interests: none of the variable of interest showed a significant Shapiro-Wilk test in any group (all ps > 0.065). Independent samples t-tests were used to test the average self-reported impulsivity (BIS-11), food craving (FCQ-T and FCQ-S), and hunger between the groups. Differences were deemed as significant when two-tailed p-value < 0.05.

Neuroimaging data were analyzed by means of the SPM-based software Conn functional connectivity toolbox (v.21a) and run on MATLAB 2016b.^30^ At second-level, we performed the following contrasts: (i) average positive and negative VTA rsFC across the groups (one-sample T-test), (ii) greater VTA rsFC in obese patients (obese > healthy weight, two-samples T-test), (iii) lower VTA rsFC in obese patients (healthy weight > obese, two-samples T-test). All the reported results survive a correction for multiple comparisons: we used the nested-taxonomy strategy recommended by Friston and colleagues, including regional effects meeting either a clusterwise or voxelwise FWER correction.^8^ The voxelwise threshold applied to the statistical maps before the clusterwise correction was p < 0.001 uncorrected, as recommended by Flandin and Friston.^6^ For clusters significant at the p < 0.05 FWER-corrected level, we also report the other peaks at p<0.001.

Brain-behavior correlations were computed, across the whole sample, between the rsFC values of eventual clusters of significant difference between the groups (i.e., by extracting the average strength of the rsFC from the cluster) and (i) impulsivity (BIS-11 scores), (ii) food craving (FCQ-T and FCQ-S scores), (iii) hunger, (iv) implicit cognitive bias (food and non-food). The Shapiro-Wilk test was significant for FCQ-S scores (W=0.94, p=0.03), hunger ratings (W=0.93, p=0.007), and D600 food scores (W=0.95, p=0.04): therefore, Spearman bivariate correlation were computed. For all the other variables of interest the Pearson correlation coefficient was computed. Correlations were deemed significant when two-tailed p < 0.05.

## 3. Results

### 3.1. Self-report data

#### Impulsivity, food craving, and hunger

Our results showed no significant difference in BIS-11, FCQ-S, and hunger scores between the groups (all ps≥0.13), both groups reporting similar levels of self-reported impulsivity, state food craving and, hunger (Table 2, Figure 1A,C-D). We observed a significant difference in FCQ-T total scores (t(44)=-2.67, p=0.01, Cohen’s d=-0.79), patients with obesity reporting higher trait (Table 2, Figure 1B) food craving.

**Table 2.**
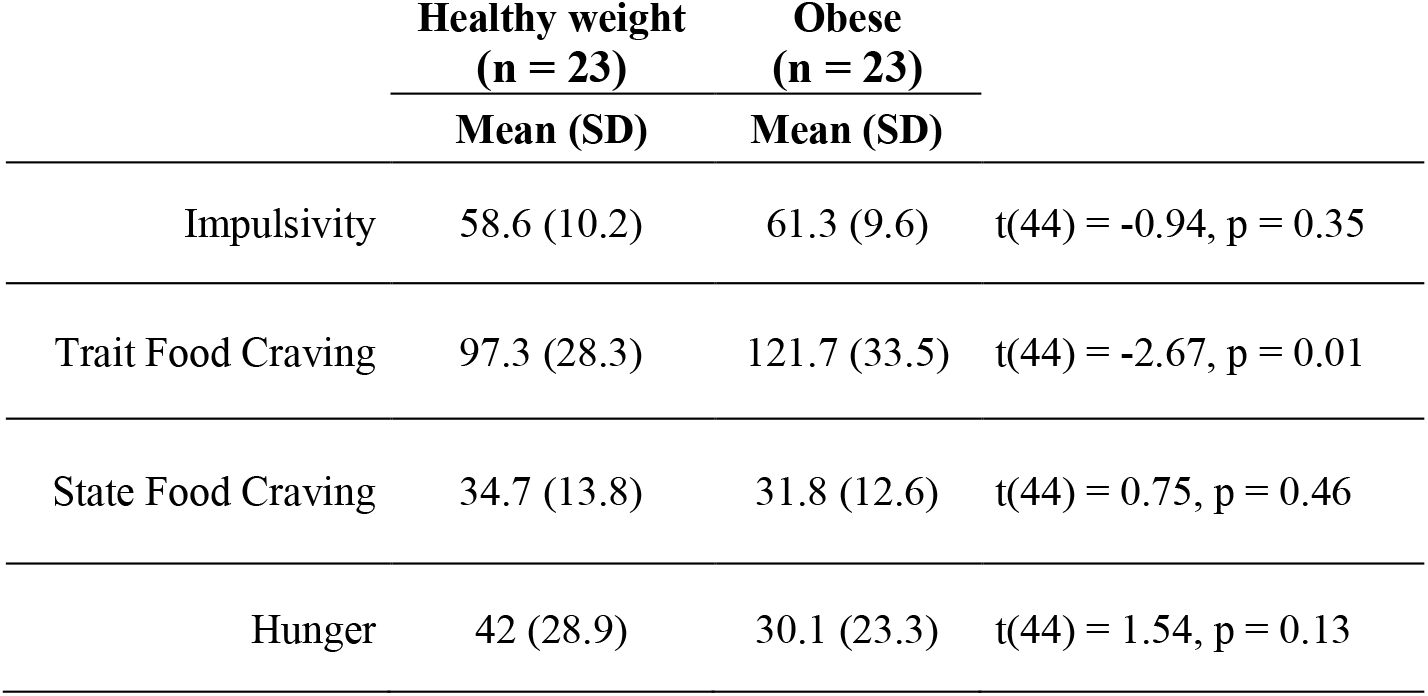
Results of the behavioral analyses.

|  | Healthy weight<br>(n = 23) | Obese<br>(n = 23) |  |
| --- | --- | --- | --- |
|  | Mean (SD) | Mean (SD) |  |
| Impulsivity | 58.6 (10.2) | 61.3 (9.6) | $t(44) = -0.94$, $p = 0.35$ |
| Trait Food Craving | 97.3 (28.3) | 121.7 (33.5) | $t(44) = -2.67$, $p = 0.01$ |
| State Food Craving | 34.7 (13.8) | 31.8 (12.6) | $t(44) = 0.75$, $p = 0.46$ |
| Hunger | 42 (28.9) | 30.1 (23.3) | $t(44) = 1.54$, $p = 0.13$ |

**Figure 1.**
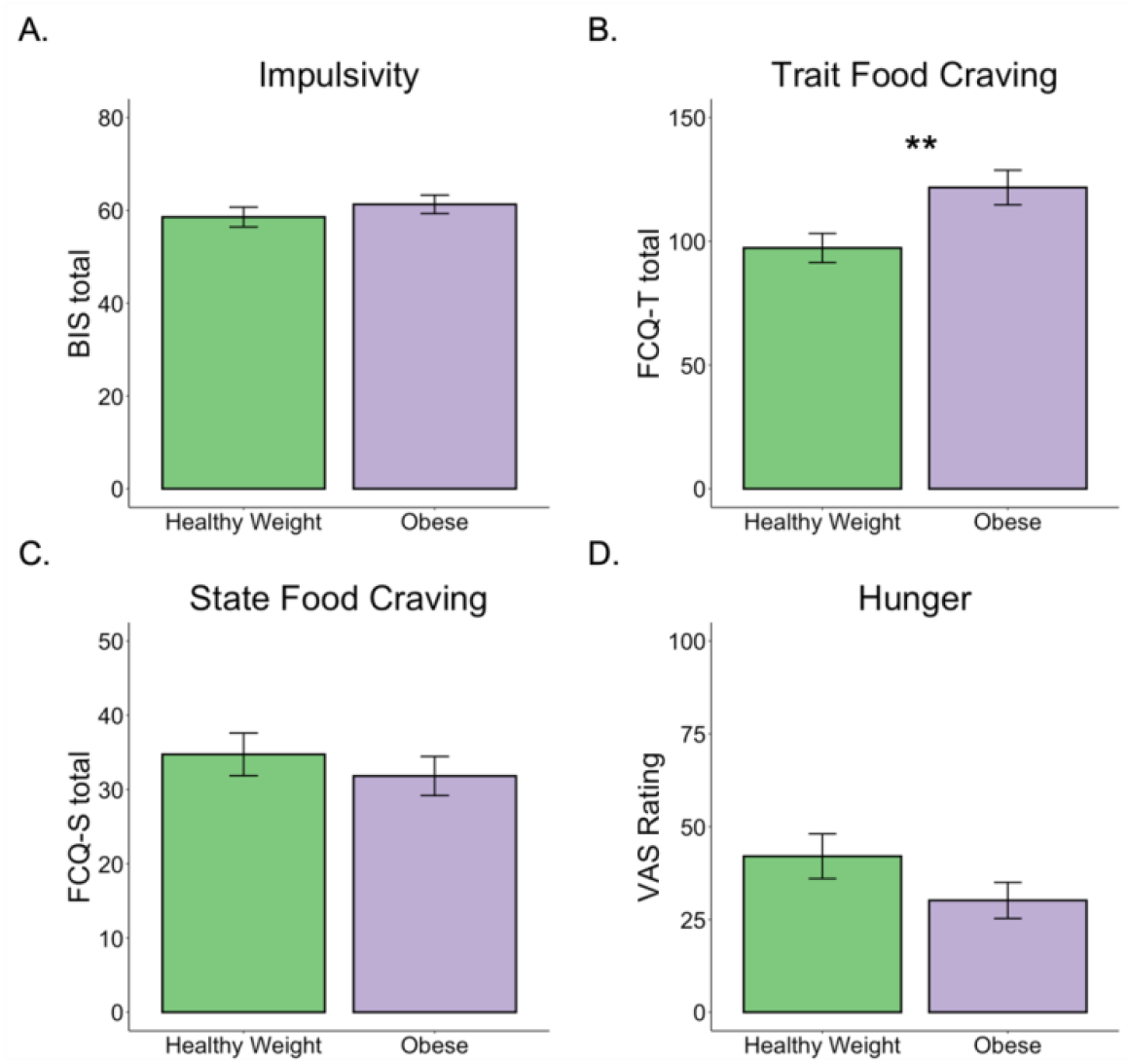
Between-group differences in behavioral measures. **A**. Impulsivity. **B**. Trait food craving. **C**. State food craving. **D**. Hunger. **, p=0.01.

### 3.2. Neuroimaging data

#### Average connectivity

##### Positive connectivity

The VTA showed average positive connectivity with a wide cortico-subcortical network including frontal (superior medial frontal gyrus, inferior frontal gyrus, supplementary motor area, anterior insula), parietal (inferior parietal lobule, angular gyrus, precuneus), occipito-temporal areas (inferior temporal gyrus, fusiform gyrus, temporal pole, lingual gyrus, cuneus, calcarine scissure), the limbic system (hippocampus, amygdala), thalamus, basal ganglia (including the nucleus accumbens, caudate, pallidum, and putamen), and the cerebellum (Table S1A and Figure S2, green).

##### Negative connectivity

The VTA showed average negative connectivity mainly with cortical areas including frontal (superior and middle frontal gyrus, precentral and postcentral gyrus, Rolandic operculum, medial orbitofrontal cortex, gyrus rectus), parietal (superior and inferior parietal lobule, precuneus), temporal (inferior and middle temporal gyrus, fusiform gyrus, parahippocampal gyrus), occipital (superior, middle, inferior occipital gyrus, lingual gyrus, cuneus), and the cerebellum (Table S1B and Figure S2, violet).

#### Obese hyper-connectivity

Compared to healthy weight participants, patients with obesity showed increased functional coupling of the bilateral VTA with the left and right ventral occipitotemporal cortex (vOTC), including the fusiform gyrus, the lingual gyrus, and extending to the cerebellum (Table 3A, Figure 2, red). The local maxima of these clusters in the fusiform gyri had stereotactic coordinates at millimetric distance from those reported for the food-selective regions of the ventral visual pathway, recently discovered by different research groups^1^.^31-33^

**Table 3.**
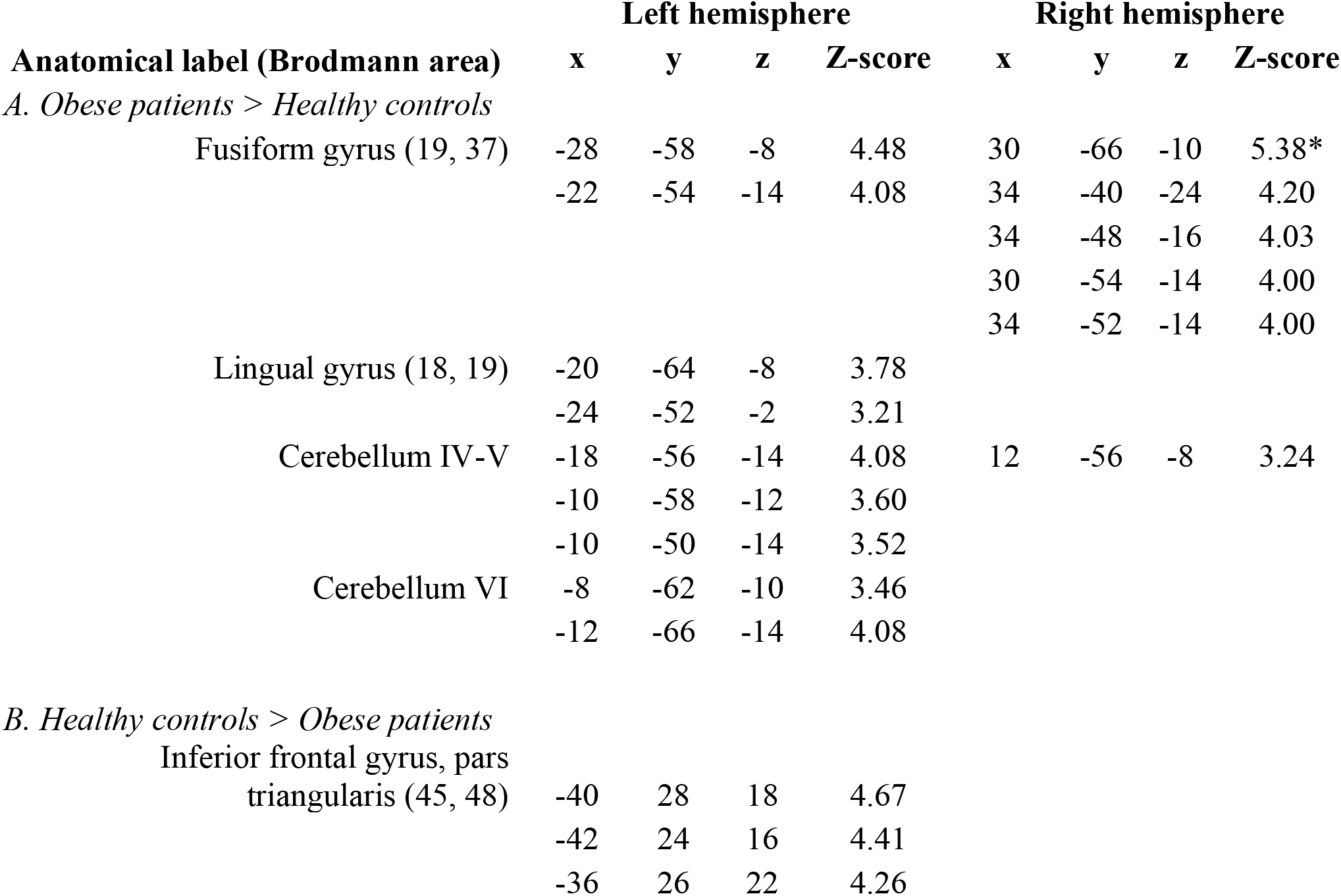
Results of the independent samples t-test on the VTA functional connectivity. **A**. Obese hyper-connectivity. **B**. Obese hypo-connectivity. For each peak, the anatomical label according with the AAL3 (Brodmann areas), the MNI coordinates, and the Z-score are reported. * p<0.05 FWE-corrected voxel-level.

**Figure 2.**
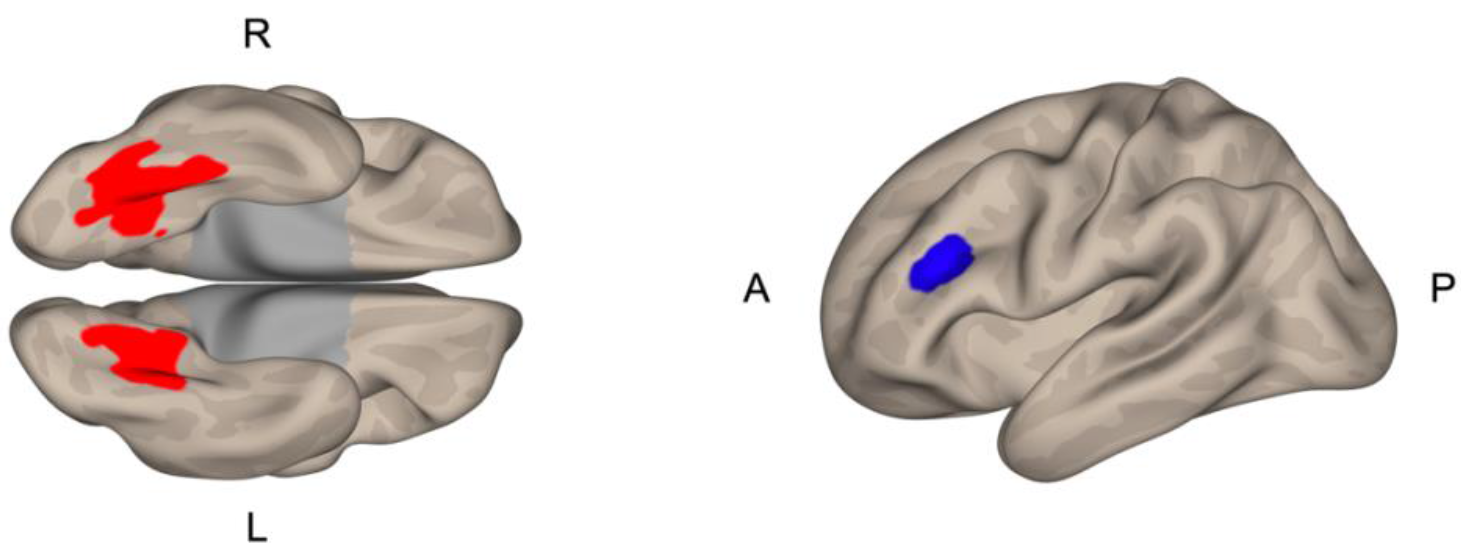
Between-group differences in VTA functional connectivity. Obese hyper-connectivity between the bilateral VTA and the left and right vOTC (left, red) and obese hypo-connectivity between the bilateral VTA and the left IFG (right, blue). IFG, inferior frontal gyrus; vOTC, ventral occipitotemporal cortex.

#### Obese hypo-connectivity

Compared to healthy weight participants, patients with obesity showed decreased functional coupling of the bilateral VTA with the left inferior frontal gyrus (IFG), pars triangularis (Table 3B, Figure 2, blue).

### 3.3. Brain-behavior correlations

#### Impulsivity, food craving, and hunger

No significant association was found between BIS-11, FCQ-S, and hunger scores and the connectivity strength of any of the clusters emerged in the univariate analyses (all ps≥0.09). FCQ-T total scores showed a significant negative association with VTA-left IFG functional connectivity (r_(46)_=-0.30, p=0.04), suggesting that higher rsFC values are associated with lower trait food craving, and a significant positive association with VTA-left vOTC (r_(46)_=0.31, p=0.03) functional connectivity, indicating that higher rsFC values are associated with higher trait food craving (Figure 3). We observed no significant association between FCQ-T total scores and VTA-right vOTC (r_(46)_=0.22, p=0.14) functional connectivity.

**Figure 3.**
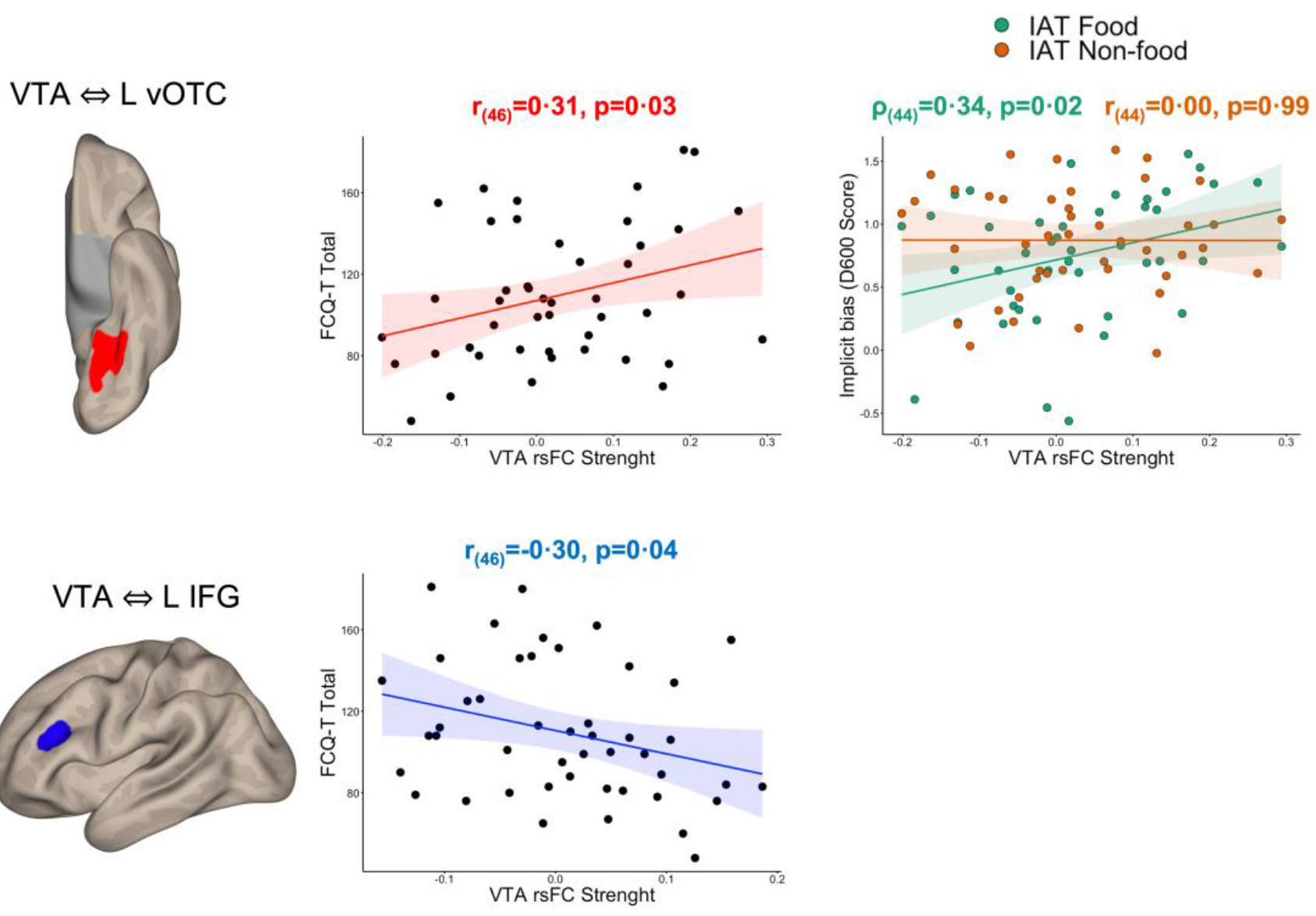
Brain-behavior correlations. Bivariate correlations between the VTA-left vOTC rsFC strength, trait food craving, and implicit bias (food, non-food; top) and between the VTA-left IFG rsFC strength and trait food craving (bottom).

#### Implicit bias for food

D600 scores of the food version of the IAT showed a significant positive association with the VTA-left vOTC functional connectivity (ρ_(44)_=0.34, p=0.02), such that the greater the functional coupling between the two regions, the greater the implicit bias for food (Figure 3). No other significant correlation with D600 food (all ps>0.24) and non-food scores (all ps>0.35) were found for any of the clusters emerged in the univariate analyses.

## 4. Discussion

This study shows for the first time that the VTA, a key structure involved in motivation and reward appreciation, has altered connectivity patterns with specific brain regions in patients with obesity: it is **hyper**-connected with the ventral visual occipito-temporal cortex (vOTC), in a region specialized in visual processing of food-related images, and **hypo**-connected with the left inferior frontal gyrus (IFG).^34^ We also showed that the connectivity strength between the VTA and these cortical regions correlates with food craving and with a bias towards high-calories foods. Notably, the same connectivity patterns do not reflect general impulsivity or psychophysiological contingencies, such as state food-craving due to starvation and appetite. These findings reveal that food-related visual stimuli become cravingly salient through an imbalanced connectivity of the reward system with sensory specific visual regions and the PFC involved in cognitive control.

As we will discuss, our results are in line with the incentive sensitization theory, applied to addiction and obesity.^18^ In what follows we provide more arguments in favour of these conclusions, and we allude at possible further directions in this research while spelling out also some limitations of the present work.

### 4.1. Complementary pathways to overeating: enhanced cue-reward associations and weak cognitive control

At the neurofunctional level, across groups, the VTA resulted functionally connected with the typical targets of its dopaminergic projections, including the thalamus, amygdala and hippocampus, the caudate nucleus, putamen, ventral striatum (extending to the nucleus accumbens), the dorsomedial PFC (dorsal ACC), and the medial orbitofrontal cortex and gyrus rectus. We also found evidence of functional coupling with regions usually not considered as part of its dopaminergic projections, including frontal (superior frontal gyrus, precentral gyrus, postcentral gyrus, anterior insula), parietal (superior and inferior parietal gyrus, supramarginal gyrus, posterior cingulate cortex, precuneus), temporal (temporal pole, inferior temporal gyrus, fusiform gyrus), occipital areas (inferior occipital and lingual gyri), and the cerebellum. Connectivity with regions outside of the mesocorticolimbic circuitry is also not surprising, once considered the neurochemical heterogeneity of neurons (dopaminergic, GABAergic, glutamatergic, combinatorial) in the ventral tegmental region, and it is largely consistent with primate anatomy and with the results of previous studies employing a seed-based approach to map the functional connections of the VTA with resting-state fMRI.^9-16^

Our main results indicate that obesity is associated with increased coupling of the VTA with the vOTC, bilaterally. Moreover, the connectivity strength of the VTA with the left vOTC was positively associated with food craving, and only with the cognitive bias for food (vs. non-food) items, suggesting that midbrain signals may enhance the incentive value of visual food representations held in the vOTC. To make this interpretation plausible one needs evidence for at least three suppositions: a role of the VTA in rewarded associative learning, beyond the basal ganglia; a specific response of discrete cortical patches for food-related visual stimuli; specific correlations of the present functional connectivity patterns with food-related behavior/psychological traits and obesity. We believe that these are all available.

First, the VTA, with a well-known role in reward processing^34^ on target neurons of the striatum, has now a recognized role in cue-reward associations within specific visual cortices: in monkeys, the presentation of unexpected rewards causes a decrease fMRI activity within cue-specific cortical regions that were paired with rewarded trials; consistent with the making of cue-reward associations, this cortical response correlated with the one measured in the VTA.^35^

Second, the vOTC, that exhibits discrete regions highly specialized in the perception of faces, bodies, scenes, or words, it also has a well-defined cortical patch exhibiting a selectivity for pictures of food^31-33^. Importantly, our local maxima fall at millimetric distance from what reported by Pennock et al. for pictures of food.^33^

Third, the relevance of the present functional brain connectivity patterns for obesity is reinforced by specific correlations with food-related behaviors and traits. The rsFC with the left vOTC was associated with the implicit bias for high-calories food, but not for control stimuli, suggesting that this pathway supports specifically the processing of food stimuli. Interestingly, the lateralization of the food-specific correlation with the implicit bias is consistent with the evidence that food-selective responses in the ventral visual pathway show a trend for a left-lateralized response^31^.

We suggest that greater connectivity between the VTA and food-selective regions of the vOTC at rest reflects stronger cue-reward associations, a sign of incentive sensitization to food cues.^17^

The lack of altered mesolimbic connectivity in obesity (e.g., with nucleus accumbens) was unexpected, once considered the literature on substance and non-substance addiction.^36,37^ A simple explanation for this discrepancy may be related to the different reinforcing potency of food compared to other non-biological rewards, including drugs of abuse.^38^ As dopamine release subsequent to food intake is lower and slower, a more extensive exposure to caloric-dense palatable foods may be required to induce significant changes within this circuitry. Such changes would be expected in more severe cases of obesity, with BMI greater or equal to 40 kg/m^2^.

In keeping with our main hypotheses, obese patients showed decreased rsFC with the prefrontal cortex, here the left inferior frontal gyrus; this was inversely associated with food craving, such that the greater the meso-prefrontal connectivity, the lower the craving for food. The lateral PFC is part of a prefrontal-striatal circuit involved in the cognitive regulation of craving, and it is thought to be involved in dietary self-regulation.^14,39^ Cognitive reappraisal strategies, including thinking about the long-term costs of eating or the benefits of not eating high-calories foods, or actively suppressing food craving, are associated with increased dorsolateral and ventrolateral PFC activity, and with a decreased reactivity of reward regions including the VTA and the ventral striatum.^40,41^ The involvement of PFC-mesostriatal circuits in the regulation of food intake is further supported by task-based fMRI studies employing food-specific go/no-go tasks to assess behavioral inhibition towards high-calories foods.^15^ When required to inhibit prepotent responses to high-calories foods, activity of prefrontal regions, including the lateral PFC, is inversely correlated to BMI.^15^ These observations are complemented by the evidence that deep rTMS targeting the PFC effectively promotes weight loss by reducing functional integration properties in visual cortices and by improving functional integration in the medial PFC and fronto-parietal network, likely reflecting a greater reliance on top-down processes.^25,42^

### 4.2. Conclusions

We unraveled, for the first time, the existence of altered mesocortical connectivity in obesity. Patients with obesity exhibit increased coupling of the VTA with regions of the ventral visual pathway specialized for the perception of food stimuli, and decreased connectivity with lateral prefrontal regions typically involved in the regulation of craving and behavioral inhibition. We also clarify that these functional anatomical pathways are specifically involved in food craving and cognition, and they are not related to impulsivity or hunger. Our findings provide two main insights: first, tighter VTA-vOTC connectivity may reflect stronger cue-reward associations in obesity, which favors food craving via the automatic activation of the rewarding properties of food; second, weaker coupling of the VTA with the lateral PFC may contribute to faulty cognitive control over food craving and behavior, due to inefficient down-regulation of the midbrain via the PFC.

Our results pave the way to novel interventions to treat obesity, providing a proof of concept that non-invasive brain neurostimulation techniques may be used to modulate midbrain activity and connectivity via the lateral PFC.

### 4.2. Limitations and future directions

We are aware that the cross-sectional nature of our design prevents us to infer whether the observed differences pre-existed the morbid obese state or whether they are induced by obesity. Future longitudinal studies may contribute to disentangle this issue. Another limitation is related to the generalizability of our findings to more severe cases of obesity, which pose unique challenges to the MRI environment in terms of physical constraints and image quality. Finally, the lack of behavioural measures specifically designed to assess approach/reactivity to food cues and behavioral inhibition did not allow us to provide a more extensive explanation of our neural findings. However, correlations with behavior and biases make these concerns less pressing.

## Data Availability

All data produced in the present study are available upon reasonable request to the authors.

## Footnotes

1 Our local maxima in the fusiform gyri (left: -28, -58, -8; right: 30, -54, -14) were at millimetric distance from those reported by an Activation Likelihood Estimation (ALE) meta-analysis of brain areas responsive to food (van der Laan et al., 2011; left: -30, -56, -8; right: 28, -56, -12) that fall within the food-selective regions identified in Jain et al. (2023) and in Pennock et al. (2023).

## References

1. (NCD-RisC) NRFC. Worldwide trends in body-mass index, underweight, overweight, and obesity from 1975 to 2016: a pooled analysis of 2416 population-based measurement studies in 128.9 million children, adolescents, and adults. Lancet. Dec 16 2017;390(10113):2627–2642. doi:10.1016/S0140-6736(17)32129-3

2. Volkow ND, Wise RA, Baler R. The dopamine motive system: implications for drug and food addiction. Nat Rev Neurosci. Nov 16 2017;18(12):741–752. doi:10.1038/nrn.2017.130

3. Berridge KC, Kringelbach ML. Pleasure systems in the brain. Neuron. May 2015;86(3):646–64. doi:10.1016/j.neuron.2015.02.018

4. Syan SK, McIntyre-Wood C, Minuzzi L, Hall G, McCabe RE, MacKillop J. Dysregulated resting state functional connectivity and obesity: A systematic review. Neurosci Biobehav Rev. 12 2021;131:270–292. doi:10.1016/j.neubiorev.2021.08.019

5. Root DH, Wang HL, Liu B, et al. Glutamate neurons are intermixed with midbrain dopamine neurons in nonhuman primates and humans. Sci Rep. 08 01 2016;6:30615. doi:10.1038/srep30615

6. Yoo JH, Zell V, Gutierrez-Reed N, et al. Ventral tegmental area glutamate neurons co-release GABA and promote positive reinforcement. Nat Commun. 12 15 2016;7:13697. doi:10.1038/ncomms13697

7. Robbins TW, Everitt BJ. Neurobehavioural mechanisms of reward and motivation. Curr Opin Neurobiol. Apr 1996;6(2):228–36. doi:10.1016/s0959-4388(96)80077-8

8. Mahler SV, Berridge KC. What and when to “want”? Amygdala-based focusing of incentive salience upon sugar and sex. Psychopharmacology (Berl). Jun 2012;221(3):407–26. doi:10.1007/s00213-011-2588-6

9. Coenen VA, Schumacher LV, Kaller C, et al. The anatomy of the human medial forebrain bundle: Ventral tegmental area connections to reward-associated subcortical and frontal lobe regions. Neuroimage Clin. 2018;18:770–783. doi:10.1016/j.nicl.2018.03.019

10. Baler RD, Volkow ND. Drug addiction: the neurobiology of disrupted self-control. Trends Mol Med. Dec 2006;12(12):559–66. doi:10.1016/j.molmed.2006.10.005

11. Tomasi D, Volkow ND. Functional connectivity of substantia nigra and ventral tegmental area: maturation during adolescence and effects of ADHD. Cereb Cortex. Apr 2014;24(4):935–44. doi:10.1093/cercor/bhs382

12. Stoeckel LE, Weller RE, Cook EW, Twieg DB, Knowlton RC, Cox JE. Widespread reward-system activation in obese women in response to pictures of high-calorie foods. Neuroimage. Jun 2008;41(2):636–47. doi:10.1016/j.neuroimage.2008.02.031

13. Devoto F, Zapparoli L, Bonandrini R, et al. Hungry brains: A meta-analytical review of brain activation imaging studies on food perception and appetite in obese individuals. Neurosci Biobehav Rev. 11 2018;94:271–285. doi:10.1016/j.neubiorev.2018.07.017

14. Kober H, Mende-Siedlecki P, Kross EF, et al. Prefrontal-striatal pathway underlies cognitive regulation of craving. Proc Natl Acad Sci U S A. Aug 2010;107(33):14811–6. doi:10.1073/pnas.1007779107

15. He Q, Huang X, Zhang S, Turel O, Ma L, Bechara A. Dynamic Causal Modeling of Insular, Striatal, and Prefrontal Cortex Activities During a Food-Specific Go/NoGo Task. Biol Psychiatry Cogn Neurosci Neuroimaging. Dec 2019;4(12):1080–1089. doi:10.1016/j.bpsc.2018.12.005

16. Wang GJ, Volkow ND, Fowler JS. The role of dopamine in motivation for food in humans: implications for obesity. Expert Opin Ther Targets. Oct 2002;6(5):601–9. doi:10.1517/14728222.6.5.601

17. Robinson TE, Berridge KC. The neural basis of drug craving: an incentive-sensitization theory of addiction. Brain Res Brain Res Rev. 1993 Sep-Dec 1993;18(3):247–91.

18. Morales I, Berridge KC. ‘Liking’ and ‘wanting’ in eating and food reward: Brain mechanisms and clinical implications. Physiol Behav. 12 01 2020;227:113152. doi:10.1016/j.physbeh.2020.113152

19. 1. Zapparoli L, Devoto F, Giannini G, Zonca S, Gallo F, Paulesu E. Neural structural abnormalities behind altered brain activation in obesity: Evidence from meta-analyses of brain activation and morphometric data. Neuroimage Clin. Sep 05 2022;36:103179. doi:10.1016/j.nicl.2022.103179

20. Lips MA, Wijngaarden MA, van der Grond J, et al. Resting-state functional connectivity of brain regions involved in cognitive control, motivation, and reward is enhanced in obese females. Am J Clin Nutr. Aug 2014;100(2):524–31. doi:10.3945/ajcn.113.080671

21. Kullmann S, Heni M, Veit R, et al. The obese brain: association of body mass index and insulin sensitivity with resting state network functional connectivity. Hum Brain Mapp. May 2012;33(5):1052–61. doi:10.1002/hbm.21268

22. Wijngaarden MA, Veer IM, Rombouts SA, et al. Obesity is marked by distinct functional connectivity in brain networks involved in food reward and salience. Behav Brain Res. 2015;287:127–34. doi:10.1016/j.bbr.2015.03.016

23. Greenwald AG, McGhee DE, Schwartz JL. Measuring individual differences in implicit cognition: the implicit association test. J Pers Soc Psychol. Jun 1998;74(6):1464–80. doi:10.1037//0022-3514.74.6.1464

24. Ferrulli A, Macrì C, Terruzzi I, et al. Weight loss induced by deep transcranial magnetic stimulation in obesity: A randomized, double-blind, sham-controlled study. Diabetes Obes Metab. 08 2019;21(8):1849–1860. doi:10.1111/dom.13741

25. Devoto F, Ferrulli A, Zapparoli L, et al. Repetitive deep TMS for the reduction of body weight: Bimodal effect on the functional brain connectivity in “diabesity”. Nutr Metab Cardiovasc Dis. 06 07 2021;31(6):1860–1870. doi:10.1016/j.numecd.2021.02.015

26. Patton JH, Stanford MS, Barratt ES. Factor structure of the Barratt impulsiveness scale. J Clin Psychol. Nov 1995;51(6):768–74. doi:10.1002/1097-4679(199511)51:6<768::aid-jclp2270510607>3.0.co;2-1

27. Innamorati M, Imperatori C, Balsamo M, et al. Food Cravings Questionnaire-Trait (FCQ-T) discriminates between obese and overweight patients with and without binge eating tendencies: the Italian version of the FCQ-T. J Pers Assess. 2014;96(6):632–9. doi:10.1080/00223891.2014.909449

28. Greenwald AG, Nosek BA, Banaji MR. Understanding and using the implicit association test: I. An improved scoring algorithm.. Journal of personality and social psychology,. 2003;85(2):197.

29. Rorden C, Brett M. Stereotaxic display of brain lesions. Behav Neurol. 2000;12(4):191–200.

30. Whitfield-Gabrieli S, Nieto-Castanon A. Conn: a functional connectivity toolbox for correlated and anticorrelated brain networks. Brain Connect. 2012;2(3):125–41. doi:10.1089/brain.2012.0073

31. Khosla M, Ratan Murty NA, Kanwisher N. A highly selective response to food in human visual cortex revealed by hypothesis-free voxel decomposition. Curr Biol. Oct 10 2022;32(19):4159-4171.e9. doi:10.1016/j.cub.2022.08.009

32. Jain N, Wang A, Henderson MM, et al. Selectivity for food in human ventral visual cortex. Commun Biol. Feb 15 2023;6(1):175. doi:10.1038/s42003-023-04546-2

33. Pennock IML, Racey C, Allen EJ, et al. Color-biased regions in the ventral visual pathway are food selective. Curr Biol. Jan 09 2023;33(1):134-146.e4. doi:10.1016/j.cub.2022.11.063

34. Schultz W, Dayan P, Montague PR. A neural substrate of prediction and reward. Science. Mar 1997;275(5306):1593–9.

35. Arsenault JT, Nelissen K, Jarraya B, Vanduffel W. Dopaminergic reward signals selectively decrease fMRI activity in primate visual cortex. Neuron. Mar 2013;77(6):1174–86. doi:10.1016/j.neuron.2013.01.008

36. Gu H, Salmeron BJ, Ross TJ, et al. Mesocorticolimbic circuits are impaired in chronic cocaine users as demonstrated by resting-state functional connectivity. Neuroimage. Nov 2010;53(2):593–601. doi:10.1016/j.neuroimage.2010.06.066

37. Wang R, Li M, Zhao M, et al. Internet gaming disorder: deficits in functional and structural connectivity in the ventral tegmental area-Accumbens pathway. Brain Imaging Behav. Aug 2019;13(4):1172–1181. doi:10.1007/s11682-018-9929-6

38. Volkow ND, Wise RA. How can drug addiction help us understand obesity? Nat Neurosci. May 2005;8(5):555–60. doi:10.1038/nn1452

39. Lowe CJ, Reichelt AC, Hall PA. The Prefrontal Cortex and Obesity: A Health Neuroscience Perspective. Trends Cogn Sci. Apr 2019;23(4):349–361. doi:10.1016/j.tics.2019.01.005

40. Yokum S, Stice E. Cognitive regulation of food craving: effects of three cognitive reappraisal strategies on neural response to palatable foods. Int J Obes (Lond). Dec 2013;37(12):1565–70. doi:10.1038/ijo.2013.39

41. Demos McDermott KE, Lillis J, McCaffery JM, Wing RR. Effects of Cognitive Strategies on Neural Food Cue Reactivity in Adults with Overweight/Obesity. Obesity (Silver Spring). Oct 2019;27(10):1577–1583. doi:10.1002/oby.22572

42. Kim SH, Park BY, Byeon K, et al. The effects of high-frequency repetitive transcranial magnetic stimulation on resting-state functional connectivity in obese adults. Diabetes Obes Metab. 08 2019;21(8):1956–1966. doi:10.1111/dom.13763

