## Supplementary materials for "How Images of Food Become Cravingly Salient in Obesity"

**1. Supplementary Methods**

### 1. Supplementary methods
#### 1.1. Implicit Association Test
##### 1.1.1. Stimuli selection
Before the experimental session, each participant was administered an online inventory that comprises a list of 100 food stimuli (50 high-calories (HC), palatable foods and 50 low-calories (LC) foods) presented in a fixed random order. Participants rated each food in the list on a 6-point Likert scale [“How much is this food palatable for you”; 1 = not palatable at all, 6 = extremely palatable]. For each participant, we randomly selected 6 food pictures among the most liked (liking rating = 5-6) HC foods and 6 food pictures among the least liked (liking rating = 1-2) LC stimuli that were included in the food version of the Implicit Association Test (IAT Food).[1] In the control version of the IAT (Non-food), which contrasted pictures of weapons and pictures of flowers, the visual stimuli were the same for all the participants. The purpose of the non-food version of the IAT was to control for the general cognitive processes required to categorize food-unrelated items belonging to opposite categories.

Food stimuli were taken from the Food-pics database, a standardized and validated database of food pictures differing in caloric and macronutrient content.[2] HC food stimuli included caloric-dense and fat foods such as pizza, French fries, and chocolate snacks, whereas LC foods mainly consisted of vegetables (e.g., spinaches, salad, carrots) and low-fat foods rich in proteins (e.g., fish, grilled chicken meat). Word stimuli were taken from a previous study employing an Italian version of the IAT: hence, 6 words with markedly positive valence – love (amore), joy (gioia), friend (amico), kiss (bacio), hug (abbraccio), peace (pace) – and 6 words with markedly negative valence – killer (assassin), cadaver (cadavere), torture (tortura), death (morte), agony (agonia), murder (omicidio) – were used as word stimuli in the two versions of the IAT.[3]

The caloric content of food stimuli was never explicitly mentioned throughout the experimental session, nor the association between caloric content and liking was made explicitly available to them.

##### 1.1.2. Task description
The IAT was used to assess implicit processing of food-related (food: HC and LC foods) and food-unrelated (non-food: flowers and weapons) stimuli.[1] Each version of the IAT consisted of seven blocks, as indicated by Greenwald and colleagues, during which participants had to classify: in block 1, participants were asked to classify *palatable* (left key) and *unpalatable* (right key) food pictures. In block 2, participants were asked to classify *positive* (left key) and *negative* (right key) words. In blocks 3 and 4 (congruent condition; Figure S1, top left panel), both food pictures and word stimuli were randomly presented, one item at the time, and participants were asked to classify *palatable* food pictures and *positive* words (left key) and *unpalatable* food pictures and *negative* words (right key). In block 5, the category-response mapping of block 1 were reversed: participants were asked to sort *unpalatable* food pictures (left key) and *palatable* food pictures (right key). Finally, in blocks 6 and 7 (incongruent condition; Figure S1, bottom left panel), both food pictures and word stimuli were randomly presented, one item at a time, and participants were asked to sort *unpalatable* food pictures and *positive* words (left key) and *palatable* food pictures and *negative* words (right key).[4]

The same scheme of block presentation was used for the non-food version of the IAT, in which palatable and unpalatable food pictures were replaced by *flower* and *weapons* pictures (Figure S1, top and bottom right panels). Half of the participants completed the task with the congruent condition first: for the remaining participants (incongruent condition first), the blocks 1, 3, and 4 were switched with blocks 5, 6, and 7, respectively.[4] Participants classified the stimuli by pressing a button with their left or right index finger. The labels for food stimuli were “palatable” and “unpalatable”, whereas for non-food stimuli were “flowers” and “weapons”. The labels for positive and negative words were “positive” and “negative”, respectively. Labels were displayed on the top left and right corners of the screen and were visible throughout the task. Participants were instructed to respond as quickly as possible: reaction times and response accuracy were recorded to compute the D600 score for each version of the task. The order of the IATs (food, non-food) was counterbalanced across subjects. Each stimulus (picture or word) was displayed for 3000 ms and with a random inter-trial interval (randomized ITI = 1000 – 3000 ms; average: 2000 ms). Each version of the task lasted for approximately 15 minutes.

### 1.2. Neuroimaging

#### 1.2.1. fMRI data acquisition

Whole-brain functional images were obtained with a T2*-weighted gradient-echo, EPI pulse sequence, using BOLD contrast (flip angle = 90°, TE = 60 ms, TR = 2000 ms, FOV = 250 mm and matrix = 64 x 64, slice thickness was 4 mm). We acquired one functional run comprising 350 scans (duration: ≃ 12 minutes). The functional run of one obese patient was interrupted ~1 minute earlier due to physical discomfort during data acquisition (n=316 scans); as removing the patient from the analyses did not change the fMRI findings, the subject was included in the final sample. Subjects were instructed to lay with their eyes closed without focusing on anything in particular. Fifteen initial "dummy" scans were acquired but discarded before fMRI analysis. All subjects were also scanned with an MPRAGE high-resolution T1-weighted volumetric scan for further visualization of the results (flip angle = 35°, TE = 5 ms, TR = 21 ms, FOV = 250 mm, matrix = 256 x 256, TI = 768, for a total of 160 slices with 1 x 1 x 1 mm voxels).

#### 1.2.2. Preprocessing

Preprocessing and subsequent analysis of fMRI data were performed with the SPM-based software Conn functional connectivity toolbox (version 21a), and run on MATLAB (version 2016b; The MathWorks, Inc., Natick, Massachusetts, United States).[5] We run the "standard" preprocessing pipeline available in Conn, which included: slice-time correction, realignment of the functional images to the first image of the series, T1-weighted image segmentation, co-registration of the mean functional image to the structural image, spatial normalization to the Montreal Neurological Institute (MNI) template and spatial smoothing. The slice-time corrected, realigned, co-registered and normalized images were smoothed with a 10x10x10mm full-width half-maximum (FWHM) isotropic Gaussian kernel. This level of smoothing is recommended for the use of Gaussian random fields theory to perform cluster level correction for multiple comparisons.[6] Scan outliers based on inter-scan global signal changes and movement were detected with the Artifact Detection Tools (ART, http://www.nitrc.org/projects/artifact_detect). Scans were considered as outlier when the scan-to-scan global signal difference exceeded 2 standard deviations of the mean, and when the compounded measure of movement parameters exceeded 2 mm scan-to-scan movement.
Then, functional images were band-pass filtered to 0.008~0.09 Hz (the frequency range of spontaneous BOLD signal fluctuations) and motion regressed (six motion parameters included as a first-level covariate) to reduce the influence of noise. To regress-out the BOLD signal associated with the individual white matter (WM) and cerebrospinal fluid (CSF), the WM and CSF masks were entered as confound, following the CompCor strategy implemented in Conn.[7] Additional preprocessing steps included linear detrending and despiking (after regression) of the time-series.

### 2. Supplementary tables

#### 2.1. Table S1

**Table S1 | Results of the one-sample t-tests.** Average positive and negative VTA functional connectivity across the whole sample. For each peak, the anatomical label according with the AAL3 (Brodmann areas), the MNI coordinates, and the Z-score are reported. * p <0.05 FWE-corrected voxel-level.

| Anatomical label (Brodmann area) | Left hemisphere | | | | Right hemisphere | | | |
|---|---|---|---|---|---|---|---|---|
| | **x** | **y** | **z** | **Z-score** | **x** | **y** | **z** | **Z-score** |
| *A. Positive functional connectivity* | | | | | | | | |
| Superior medial frontal cortex (32) | | | | | 8 | 20 | 44 | 3.72 |
| Supplementary motor area (32) | | | | | 10 | 22 | 48 | 3.28 |
| Dorsal anterior cingulate cortex (24, 32) | -4 | 26 | 24 | 4.29 | 6 | 36 | 20 | 4.13 |
| | -6 | 30 | 22 | 4.20 | 8 | 36 | 12 | 4.05 |
| | -8 | 36 | 18 | 3.96 | 6 | 34 | 24 | 3.68 |
| | | | | | 4 | 28 | 26 | 3.51 |
| | | | | | 6 | 24 | 26 | 3.28 |
| Pregenual anterior cingulate cortex (32) | -6 | 42 | 18 | 3.89 | 8 | 38 | 16 | 4.26 |
| | | | | | 4 | 40 | 16 | 3.96 |
| Middle cingulate cortex (23, 24) | | | | | 4 | 20 | 40 | 3.61 |
| | | | | | 10 | -46 | 32 | 3.55 |
| | | | | | 2 | 26 | 36 | 3.51 |
| Inferior parietal cortex (39) | | | | | 58 | -58 | 38 | 4.18 |
| Angular gyrus (39) | | | | | 52 | -64 | 44 | 3.24 |
| Posterior cingulate cortex (23, 26) | 8 | -42 | 22 | 3.82 | | | | |
| | 8 | -48 | 26 | 3.65 | | | | |
| Precuneus (7, 23) | | | | | 2 | -70 | 38 | 4.09 |
| | | | | | 6 | -64 | 32 | 3.77 |
| | | | | | 6 | -56 | 26 | 3.74 |
| | | | | | 8 | -52 | 28 | 3.64 |
| | | | | | 6 | -62 | 28 | 3.45 |
| Superior occipital cortex (17) | | | | | 12 | -100 | 14 | 3.87 |
| Cuneus (17) | | | | | 18 | -100 | 10 | 4.05 |
| Calcarine cortex (17, 18) | | | | | 8 | -92 | 10 | 3.51 |
| | | | | | 4 | -88 | 6 | 3.17 |
| Amygdala (34) | -28 | -4 | -16 | 5.51* | | | | |
| Pallidum | -12 | -4 | -2 | 5.89* | 28 | -6 | -8 | 5.38* |
| Hippocampus (34) | | | | | 20 | -6 | -10 | 5.44* |
| Thalamus (LGN) | -22 | -22 | -8 | 6.81* | | | | |
| Thalamus (VA) | -10 | -8 | 2 | 5.68* | | | | |
| Thalamus (VL) | -10 | -10 | 6 | 5.56* | | | | |
| Ventral tegmental area | 0 | -22 | -18 | Inf* | 4 | -22 | -16 | Inf* |
| Substantia nigra (pars reticulata) | -10 | -6 | -10 | 6.88* | | | | |
| Cerebellum IV-V | -6 | -48 | -16 | 5.87* | 8 | -44 | -12 | 6.37* |
| Cerebellum VIII | -30 | -58 | -52 | 4.08 | 14 | -60 | -44 | 4.68 |

| | | | | | | | | |
|---|---|---|---|---|---|---|---|---|
| Cerebellum IX | -12 | -40 | -46 | 4.50 | 20 | -46 | -50 | 4.35 |
| | -18 | -48 | -44 | 4.02 | 14 | -56 | -48 | 4.12 |
| | | | | | 18 | -52 | -46 | 4.12 |
| | | | | | 18 | -50 | -50 | 4.02 |
| Cerebellum X | -16 | -42 | -42 | 4.45 | | | | |
| Vermis III | -2 | -44 | -14 | 5.94* | | | | |
| Vermis VI | | | | | 2 | -68 | -14 | 6.27* |
| | | | | | 6 | -66 | -16 | 5.85* |
| Vermis IX | | | | | 2 | -58 | -40 | 4.20 |
| *B. Negative functional connectivity* | | | | | | | | |
| Superior frontal gyrus (6) | -24 | -2 | 74 | 3.70 | | | | |
| | -24 | -6 | 72 | 3.61 | | | | |
| | -18 | -4 | 58 | 3.39 | | | | |
| Precentral gyrus (4, 6) | -34 | -2 | 62 | 4.39 | 36 | -24 | 56 | 3.41 |
| | -32 | -6 | 60 | 4.34 | 38 | -16 | 56 | 3.21 |
| | -34 | -28 | 58 | 4.11 | | | | |
| | 36 | -24 | 56 | 3.41 | | | | |
| | 38 | -16 | 56 | 3.21 | | | | |
| Medial orbitofrontal cortex (11) | | | | | 24 | 28 | -28 | 3.77 |
| | | | | | 22 | 34 | -28 | 3.68 |
| | | | | | 14 | 38 | -26 | 3.45 |
| Rectus (11) | | | | | 10 | 40 | -28 | 3.45 |
| Postcentral gyrus (2, 3) | -28 | -34 | 68 | 4.38 | 38 | -34 | 54 | 4.68 |
| | -30 | -40 | 68 | 4.12 | 66 | -4 | 16 | 3.97 |
| | -36 | -30 | 64 | 4.03 | 62 | 2 | 14 | 3.85 |
| | -44 | -34 | 62 | 3.92 | 34 | -28 | 38 | 3.73 |
| | -42 | -38 | 62 | 3.92 | 36 | -24 | 36 | 3.52 |
| | -38 | -42 | 60 | 3.87 | 34 | -30 | 44 | 3.52 |
| Superior parietal lobule (7, 40) | -16 | -62 | 52 | 4.39 | | | | |
| | -28 | -44 | 64 | 4.22 | | | | |
| | -34 | -46 | 56 | 3.96 | | | | |
| | -18 | -58 | 54 | 3.95 | | | | |
| | -30 | -56 | 66 | 3.94 | | | | |
| | -30 | -52 | 64 | 3.89 | | | | |
| | -38 | -46 | 58 | 3.86 | | | | |
| Inferior parietal lobule (40) | -36 | -46 | 52 | 4.04 | | | | |
| | -40 | -40 | 52 | 3.94 | | | | |
| Supramarginal gyrus (40) | 38 | -32 | 34 | 4.30 | | | | |
| | 38 | -34 | 42 | 3.79 | | | | |
| Inferior temporal gyrus (20,21) | -46 | -22 | -26 | 5.84* | 52 | -38 | -20 | 4.97* |
| | -48 | -26 | -26 | 5.76* | 56 | -22 | -34 | 4.95* |
| | -40 | -26 | -24 | 5.69* | 52 | -46 | -20 | 4.64 |
| | -44 | -32 | -26 | 5.62* | 58 | -52 | -16 | 4.63 |

| | | | | | | | | |
|---|---|---|---|---|---|---|---|---|
| | -52 | -40 | -28 | 5.5* | 58 | -48 | -20 | 4.45 |
| | -50 | -30 | -24 | 5.48* | 66 | -36 | -22 | 3.92 |
| | -52 | -30 | -20 | 5.35* | 50 | -56 | -18 | 3.72 |
| | -58 | -30 | -26 | 5.29* | 46 | -70 | -12 | 3.61 |
| | -56 | -32 | -22 | 5.24* | | | | |
| | -64 | -8 | -30 | 3.92 | | | | |
| Fusiform gyrus (19,20) | | | | | 50 | -72 | -18 | 4.74 |
| | | | | | 38 | -22 | -24 | 4.07 |
| | | | | | 40 | -28 | -26 | 3.79 |
| Superior occipital gyrus (19) | | | | | 24 | -76 | 34 | 3.95 |
| | | | | | 22 | -80 | 38 | 3.74 |
| Middle occipital gyrus (19) | | | | | 30 | -76 | 20 | 4.02 |
| | | | | | 30 | -78 | 24 | 3.75 |
| | | | | | 36 | -80 | 12 | 3.49 |
| | | | | | 38 | -80 | 22 | 3.49 |
| | | | | | 36 | -82 | 16 | 3.40 |
| Lingual gyrus (18,19) | | | | | 42 | -86 | -20 | 4.95* |
| | | | | | 18 | -98 | -20 | 4.60 |
| | | | | | 36 | -92 | -18 | 4.02 |

### 3. Supplementary figures

#### 3.1. Figure S1

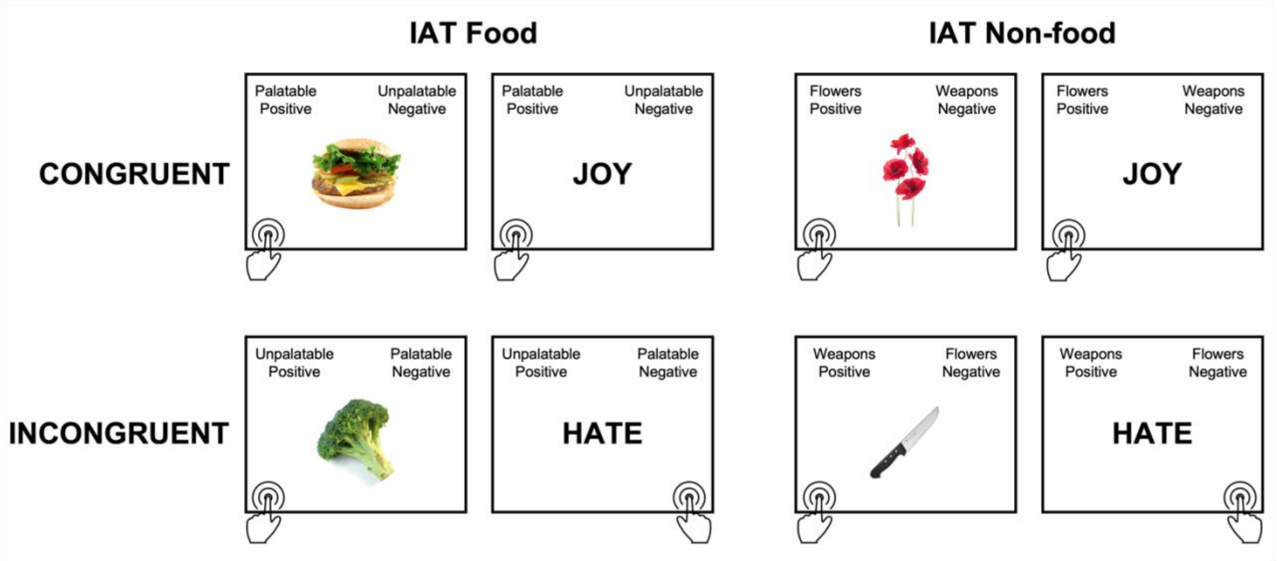

**Figure S1 | Graphical example of congruent and incongruent trials in the IAT Food and IAT Non-food.**

#### 3.2. Figure S2

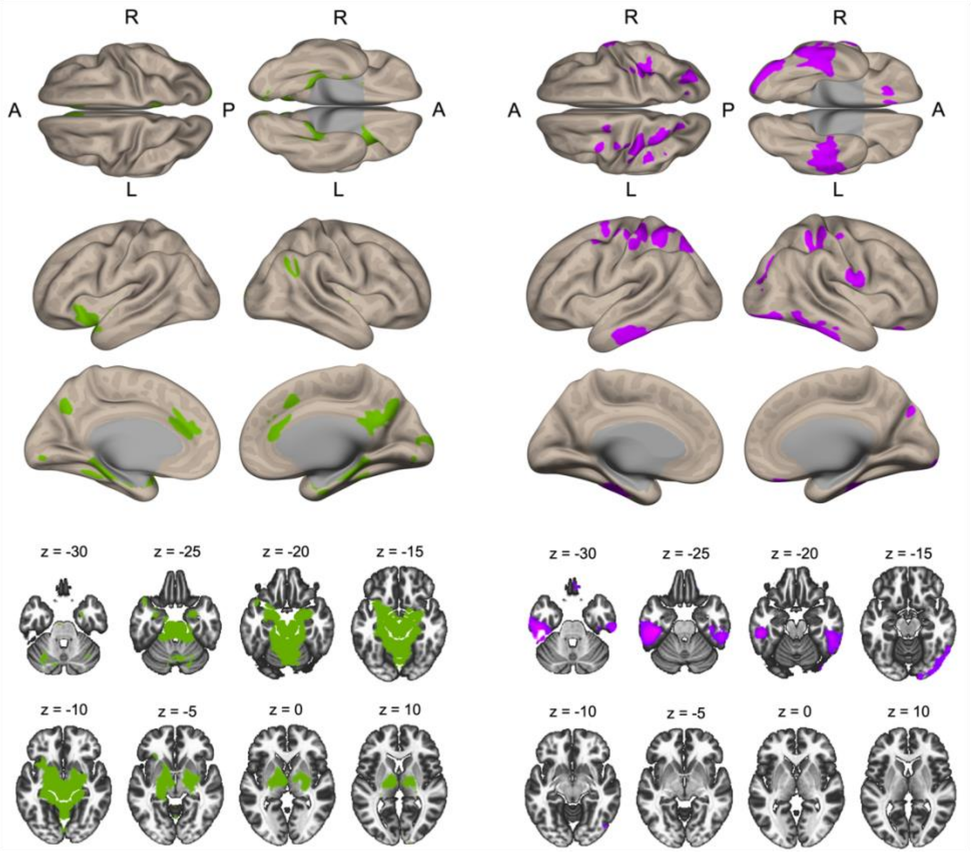

**Figure S1 | Average VTA functional connectivity in the whole sample.** Average positive (left, green) and negative (right, violet) functional connectivity of the VTA in the whole sample overlaid on a semi-unfolded rendering of the cortex (top) and on axial slices (bottom). Coordinates are shown in MNI stereotaxic space.

### 4. Supplementary references

#### 4.1. Reference list

1. Greenwald AG, McGhee DE, Schwartz JL. Measuring individual differences in implicit cognition: the implicit association test. *J Pers Soc Psychol*. Jun 1998;74(6):1464-80. doi:10.1037//0022-3514.74.6.1464
2. Blechert J, Meule A, Busch NA, Ohla K. Food-pics: an image database for experimental research on eating and appetite. *Front Psychol*. 2014;5:617. doi:10.3389/fpsyg.2014.00617
3. Mattavelli G, Zuglian P, Dabroi E, Gaslini G, Clerici M, Papagno C. Transcranial magnetic stimulation of medial prefrontal cortex modulates implicit attitudes towards food. *Appetite*. Jun 2015;89:70-6. doi:10.1016/j.appet.2015.01.014
4. Greenwald AG, Nosek BA, Banaji MR. Understanding and using the implicit association test: I. An improved scoring algorithm. . *Journal of personality and social psychology* , . 2003;*85*(2):197.
5. Whitfield-Gabrieli S, Nieto-Castanon A. Conn: a functional connectivity toolbox for correlated and anticorrelated brain networks. *Brain Connect*. 2012;2(3):125-41. doi:10.1089/brain.2012.0073
6. Flandin G, Friston KJ. Analysis of family-wise error rates in statistical parametric mapping using random field theory. *Hum Brain Mapp*. 05 2019;40(7):2052-2054. doi:10.1002/hbm.23839
7. Behzadi Y, Restom K, Liau J, Liu TT. A component based noise correction method (CompCor) for BOLD and perfusion based fMRI. *Neuroimage*. Aug 2007;37(1):90-101. doi:10.1016/j.neuroimage.2007.04.042
8. Friston KJ, Holmes A, Poline JB, Price CJ, Frith CD. Detecting activations in PET and fMRI: levels of inference and power. *Neuroimage*. Dec 1996;4(3 Pt 1):223-35. doi:10.1006/nimg.1996.0074
9. Haber SN, Fudge JL. The primate substantia nigra and VTA: integrative circuitry and function. *Crit Rev Neurobiol*. 1997;11(4):323-42. doi:10.1615/critrevneurobiol.v11.i4.40
10. Root DH, Wang HL, Liu B, et al. Glutamate neurons are intermixed with midbrain dopamine neurons in nonhuman primates and humans. *Sci Rep*. 08 01 2016;6:30615. doi:10.1038/srep30615
11. Yoo JH, Zell V, Gutierrez-Reed N, et al. Ventral tegmental area glutamate neurons co-release GABA and promote positive reinforcement. *Nat Commun*. 12 15 2016;7:13697. doi:10.1038/ncomms13697
12. Tomasi D, Volkow ND. Functional connectivity of substantia nigra and ventral tegmental area: maturation during adolescence and effects of ADHD. *Cereb Cortex*. Apr 2014;24(4):935-44. doi:10.1093/cercor/bhs382
13. Giordano GM, Stanziano M, Papa M, et al. Functional connectivity of the ventral tegmental area and avolition in subjects with schizophrenia: a resting state functional MRI study. *Eur Neuropsychopharmacol*. May 2018;28(5):589-602. doi:10.1016/j.euroneuro.2018.03.013
14. Murty VP, Shermohammed M, Smith DV, Carter RM, Huettel SA, Adcock RA. Resting state networks distinguish human ventral tegmental area from substantia nigra. *Neuroimage*. Oct 15 2014;100:580-9. doi:10.1016/j.neuroimage.2014.06.047
15. Zhang S, Hu S, Chao HH, Li CS. Resting-State Functional Connectivity of the Locus Coeruleus in Humans: In Comparison with the Ventral Tegmental Area/Substantia Nigra Pars Compacta and the Effects of Age. *Cereb Cortex*. Aug 2016;26(8):3413-27. doi:10.1093/cercor/bhv172
16. Wang R, Li M, Zhao M, et al. Internet gaming disorder: deficits in functional and structural connectivity in the ventral tegmental area-Accumbens pathway. *Brain Imaging Behav*. Aug 2019;13(4):1172-1181. doi:10.1007/s11682-018-9929-6
